# Replication of missense OTOG gene variants in a Brazilian cohort of Menière’s Disease

**DOI:** 10.1101/2025.04.26.25326273

**Authors:** Giselle Bianco-Bortoletto, Geovana Almeida Carneiro, Helena Fabbri-Scallet, Alberto M. Parra-Perez, Karen de Carvalho Lopes, Tatiana de Almeida Lima Sá Vieira, Fernando Freitas Ganança, Juan Carlos Amor-Dorado, Andres Soto-Varela, Jose A. Lopez-Escamez, Edi Lucia Sartorato

## Abstract

Ménière’s Disease (MD) is a chronic inner ear disorder defined by recurring episodes of vertigo, fluctuating sensorineural hearing loss, tinnitus, and/or fullness in the ear. Its prevalence varies by region and ethnicity, with scarce epidemiological data in Brazilian population. Although most MD cases are sporadic, Familial MD (FMD) is observed in 5% to 20% of European cases. By exome sequencing, we have found a rare missense variant in the *OTOG* gene in a Brazilian MD individual with probable European ancestry (chr11:17599671C>T), which was previously reported in a Spanish cohort. Two additional rare missense heterozygous *OTOG* variants were found in the same proband. Splice Site analysis showed that chr11:17599671C>T may lead to substantial changes generating exonic cis regulatory elements, and protein modelling revealed structural changes in the presence of chr11:17599671C>T, chr11:17576581G>C and chr11:17594108C>T, predicted to highly destabilize protein structure. These findings indicate that missense variants may have an additive effect leading to an unstable Otogelin and support *OTOG* gene as a key player in the MD pathophysiology.

## Introduction

Ménière’s Disease (MD, MIM 156000) is a rare chronic inner ear disorder, which manifests through recurrent episodes of vertigo, fluctuating sensorineural hearing loss (SNHL), tinnitus, and aural fullness. Its prevalence varies worldwide, ranging from 34 to 190 per 100,000 adults, with familial aggregation and higher occurrence among women and Caucasians (1,2). The lack of epidemiological data in the Brazilian population makes genetic studies an arduous task.

Although most MD cases are considered sporadic, Familial Meniere’s Disease (FMD) has been reported in 5% - 20% of cases of European descent (3). The current diagnostic approach relies on clinical criteria outlined by the Barany Society in 2015 international guidelines, emphasizing clinical stratification and the identification of incomplete phenotypes (1). Exome and genome sequencing studies combining gene burden tests and segregation analyses support MD as a polygenic condition (4–6). Around 15 genes have been linked to familial MD (3), including SNHL genes demonstrating autosomal dominant, recessive or digenic inheritance patterns (7,8) supporting genetic heterogeneity.

The *OTOG* gene, encoding Otogelin, an extracellular protein in the tectorial membrane in the organ of Corti (9) is emerging as a key gene in familial MD (10). Several missense variants in *OTOG* have been identified Spanish families with MD, showing aggregation in the coding sequence of constrained regions, that may explain higher MD prevalence in Europeans (7,10). This exome sequencing study aims to identify novel candidate genes in Brazilian patients with MD. Our results replicate a rare missense variant chr11:17599671C>T in the *OTOG* gene previously reported in a Spanish cohort in a Brazilian MD patient, supporting the role of OTOG in the molecular pathophysiology of MD.

## Materials and Methods

### Hearing Profile

Audiograms from the worse ear were analysed in *R Studio* (v.4.4.2) using regression to estimate hearing onset and outcome. Coefficient of determination (*R2*) and p-values validated the model.

### Whole Exome Sequencing (WES)

WES was performed on DNA from 23 unrelated Brazilian Menière’s disease (MD) patients. Libraries were prepared with *Nextera Rapid Exome Enrichment Kit*® (Illumina, Inc., San Diego, CA, USA) and sequenced (2×100 base pairs paired end, ≥100× coverage) on *HiSeq™ 2500* (Illumina). Reads were aligned to the GRCh38/hg38 using Burrows-Wheeler Aligner (BWA), variants were called with DeepVariant and filtered for minor allele frequency (MAF) <0.001 (Genome Aggregation Database, gnomAD v4.1.0). The Integrative Genomics Viewer (IGV) assessed read quality.

### Rare Variant Identification

Rare variants were retrieved from previously published sporadic MD (SMD, n=511) (10) and familial MD (FMD, n=100) (4) datasets (non-Finnish European MAF <0.001, CADD score >20). Pathogenicity was assessed per American College of Medical Genetics and Genomics (ACMG) and Association for Molecular Pathology (AMP) guidelines (11). Missense variant density in the *OTOG* coding sequence was determined using 200 base pair (bp) sliding windows template previously reported (10), identifying high-density regions (HDR) and low-density regions (LDR), the latter also called constraint coding regions (CCR), per gnomAD v2.1,.

### Splice Site Prediction

Human Splice Finder Pro (12) predicted donor/acceptor sites, exonic and intronic splicing enhancers (ESE/ISE), exonic and intronic splicing silencers (ESS/ISS), and branch points.

### Protein Modelling

Canonical UniProt sequences were modelled using Modeller (13) v10.5 through homology-based reconstruction using a template of the WT protein previously reported (10). Model validation included Discrete Optimized Protein Energy (DOPE), Genetic Algorithm 341 (GA341), and molecular probability density function (molpdf). Additional validation was performed via the Structural Analysis and Verification Server (SAVES v6.1; https://saves.mbi.ucla.edu/) for stereochemical quality and atomic interactions (14,15). Structural comparisons were conducted in PyMOL v3.0.

DynaMut2 (16) and MuPro (17) assessed variant impact on protein stability (ΔΔG). MuPro confidence score (CS) ranged from -1 to 1, with CS<0 indicating destabilization and CS>0 indicating stabilization. DynaMut2 classified variants as destabilizing for negative predicted free energy change (ΔΔG_pred) values and stabilizing for positive ΔΔG_pred values.

## Results

### Clinical Description

The study focuses on a male proband who meets definitive MD diagnostic criteria, with a 13-year disease course characterised by recurrent vertigo attacks (25-30 episodes), persistent left-sided tinnitus, fluctuating SNHL, and aural fullness. A comprehensive neurotological evaluation, incorporating video head impulse testing (vHIT) and serial audiometry was conducted, which confirmed bilateral SNHL without vestibular hypofunction. His medical history included well-controlled hypertension and autoimmune disease, with no familial history of auditory or vestibular disorders.

### Hearing Profile

The hearing profile of the Brazilian proband and three Spanish patients carrying the *OTOG* variant chr11:17599671C>T showed no significant hearing loss progression at any frequency (Supplementary Figure S1; Supplementary Table S1).

### Rare OTOG variant replicated in Brazilian sample

The missense variant chr11:17599671C>T (NFE MAF=0.005) was replicated in both cohorts. Two additional rare *OTOG* missense variants were identified in the Brazilian proband, chr11:17576581G>C (NFE MAF=0.00001) and chr11:17594108C>T (NFE

MAF=0.0004). All were classified as Likely pathogenic per ACMG guidelines (Table1) and located in biologically constraint low-density regions (LDR), suggesting low protein tolerance to variation (Figure 1).

**Table 1.** *OTOG* missense variants found in the proband. Chr11:17599671C>T was replicated in the Spanish cohort. For the three variants together in Otogelin DynaMut2 stability prediction was -1.9kcal/mol, destabilizing the protein.

| Variant | ID | Consequence | Amino acid change | CADD | gnomADg_AF | CSVS_AF | ABraOM_AF | ACMG* | DynaMut2 |  | MuPro |  |
| --- | --- | --- | --- | --- | --- | --- | --- | --- | --- | --- | --- | --- |
|  |  |  |  |  |  |  |  |  | Score (kcal/mol) | Prediction | Score (kcal/mol) | Prediction |
| chr11:17599671C>T | rs117005078 | Missense | p.Pro1240Leu | 33.00 | 0.005990 | 0 | 0.003416 | Likely Pathogenic | -0.49 | Destabilize | -0.24 | Decrease stability |
| chr11:17576581G>T | rs61734214 | Missense | p.Gly850Arg | 27.6 | 0.00001918 | 0 | 0.005978 | Likely Pathogenic | -0.44 | Destabilize | -0.76 | Decrease stability |
| chr11:17594108 C>T | rs7936354 | Missense | p.Pro1129Leu | 24.1 | 0.0004865 | 0 | 0.034586 | Likely Pathogenic | -0.7 | Destabilize | 0.12 | Increase stability |

\*ACMG criteria according to Oza et al. (11)

**Figure 1.**
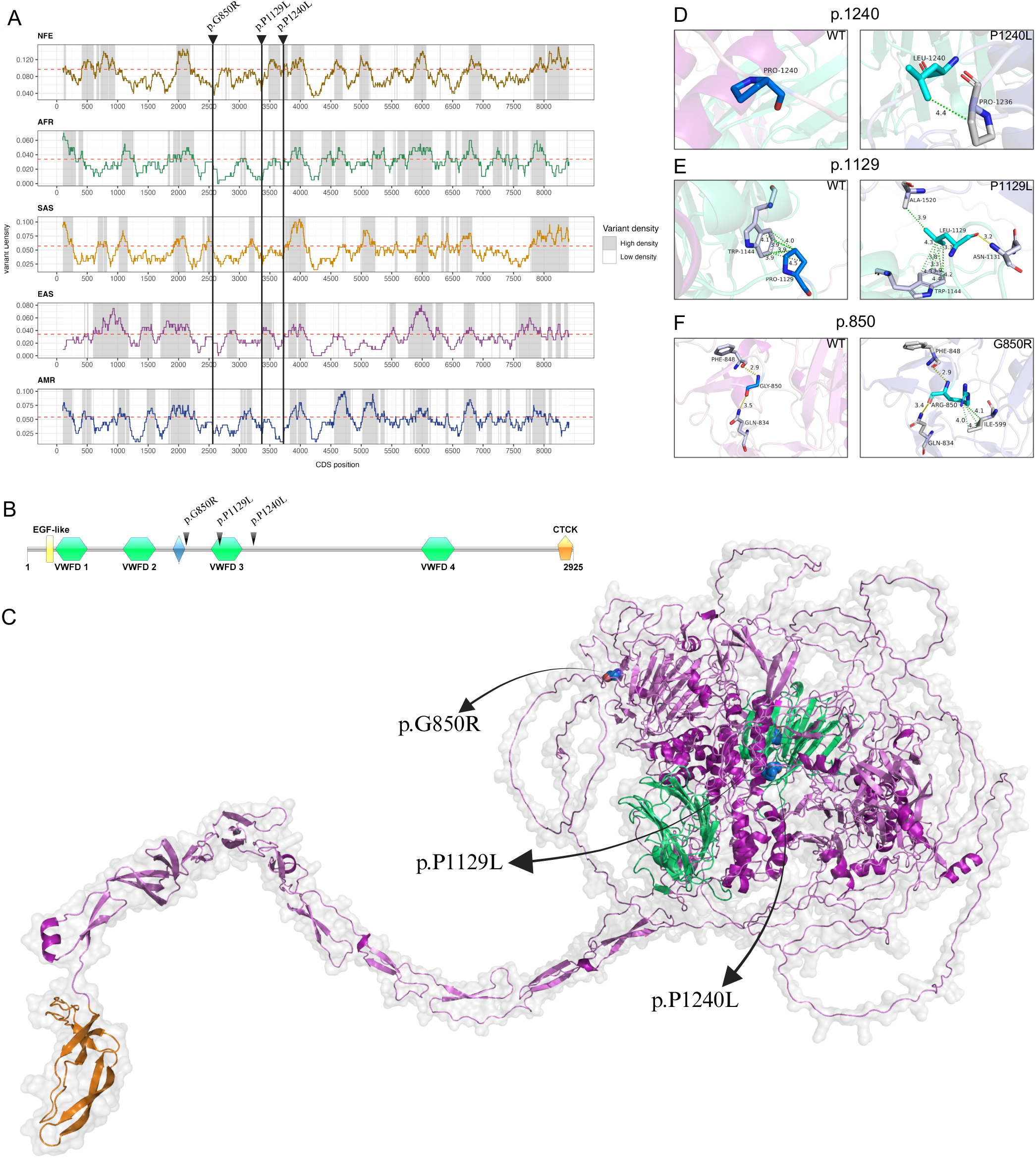
**A**. Variant density profile along the *OTOG* CDS in the NFE, AFR, SAS, EAS, and AMR populations calculated with a 201 bp sliding window. **B**. Otogelin protein domains (created with IBS software). **C**. Modelled Otogelin protein showing the three selected variants. **D-F**. Structural differences between wild type (WT) and mutated Otogelin protein at positions p.850, p.1129 and p.1240.

### Splice Site Prediction

Using HSF Pro, analysis of the three *OTOG* variants showed significant exonic cis-element alterations for chr11:17599671C>T, including the disruption of five ESEs, as well as the formation of two ESE and seven ESSs (Supplementary Table S2).

### Protein Modelling

*In silico* analysis revealed that both wild-type and mutated amino acids are in loop regions. Variant chr11:17599671C>T (p.P1240L) was predicted to not form any contacts in the WT structure, however, the mutated amino acid formed a hydrophobic contact with the Proline at p.1236 (Figure 1D).

For chr11:17594108 C>T (p.P1129L), WT Proline forms several hydrophobic contacts with p.W1144, while the variant Leucine creates new hydrophobic and polar contacts with p.W1144, p.A1520 and p.N1131 (Figure 1E).

Variant chr11:17576581G>T (p.G850R) replaced Glycine with Arginine at position 850, which is predicted to create three hydrophobic contacts with p.I599 (Figure 1F) and altering polar interactions, leading to a short α-helix at p.841-843 (Supplementary Figure S2).

Variants chr11:17599671C>T and chr11:17576581G>T were predicted to destabilize the protein in both *in silico* methods employed, while chr11:17594108 C>T showed conflicting results. When analysed together, mimicking the proband’s protein, DynaMut2 predicted high destabilization values (−1.9kcal/mol) (Supplementary Table S3).

## Discussion

This study replicates a missense variant in a Brazilian patient with SMD previously reported in 2 individuals and 1 family with MD in Spain.

There is an overload of missense variants in *OTOG* gene in FMD (10). Variant chr11:17599671C>T has been associated with moderate-to-severe flat hearing loss (60 dB) from early onset, involving all frequencies with minor low-frequency variations over time. Interestingly, it has not been reported in non-European individuals. The Brazilian proband, a male with no MD family history and unknown European ancestry, was screened for additional *OTOG* variants. Two rare heterozygous missense variants chr11:17599671C>T and chr11:17594108 C>T were identified, neither previously associated with disease.

The tectorial membrane (TM), an acellular layer above the organ of Corti, plays a key role in sound transmission by enhancing cochlear feedback. Structurally, it consists of collagen, non-glycoproteins, and proteoglycans, with Otogelin, encoded by *OTOG*, as a key non-collagenous glycoprotein. *OTOG* knockout mice have revealed its importance in auditory and balance functions, showing phenotypes like detachment of otoconial membranes, structural defects in the TM’s fibrillar network, and reduced TM resistance to sound stimuli (18). Otogelin stabilizes the TM by interacting with its constituent fibres, and *OTOG* mutations can cause moderate nonsyndromic SNHL (19).

Protein mutations alter bonding networks, affecting structure and function (20). Variants chr11:17599671C>T and chr11:17576581G>T are near, and chr11:17594108 C>T is within, the von Willebrand factor D (VWFD) domain in the Otogelin protein, which binds Ca^2+^ ions as a reservoir (3). Rare variations in Otogelin may alter electrostatic interactions and 3D structure (7).

Our study has a limitation, since we could not obtain DNA samples from the proband’s relatives for segregation analysis, but the finding of 3 rare variants in the *OTOG* gene in the same proband cannot be explained by chance.

## Conclusion

We have found a MD disease patient with 3 rare missense variants in the *OTOG* gene, which were predicted to destabilize the protein and alter its structure. This is the first time the variant chr11:17599671C>T has been reported in non-Spanish MD patients. These results support *OTOG* gene as a key player in the MD pathophysiology.

## Data Availability

The *OTOG* variants chr11:17599671C>T, chr11:17576581G>T and chr11:17594108 C>T have been deposited in ClinVar (SUB15056944. accession number pending).

## Acknowledgments

We would like to acknowledge the patient for donating blood to contribute to this study.

We thank members of CQMED-UNICAMP for their support. We thank the staff of the Life Sciences Core Facility (LaCTAD) at UNICAMP for the Genomics analysis.

## Author Contribution

Giselle Bianco-Bortoletto: Conceptualization, formal analysis, data curation, investigation, methodology, validation, visualization, writing – original draft.

Geovana Almeida Carneiro: Formal analysis, methodology.

Helena Fabbri-Scallet: Data curation.

Alberto Parra-Perez, Karen de Carvalho Lopes, Tatiana de Almeida Lima Sá Vieira, Fernando Ganança, Juan Carlos Amor-Dorado, Andres Soto-Varela: Resources.

Karen de Carvalho Lopes, Tatiana de Almeida Lima Sá Vieira: Writing – original draft.

Jose A. Lopez-Escamez, Edi Lucia Sartorato: Conceptualization, funding acquisition, project administration, resources, supervision, validation, writing – review & editing.

## Ethics

Approved by the University of Campinas School of Medical Sciences Ethics Committee (CAAE: 82809524.0.0000.5404); patient consent obtained.

