## Supplementary Figure S1 for "Replication of missense OTOG gene variants in a Brazilian cohort of Menière’s Disease"

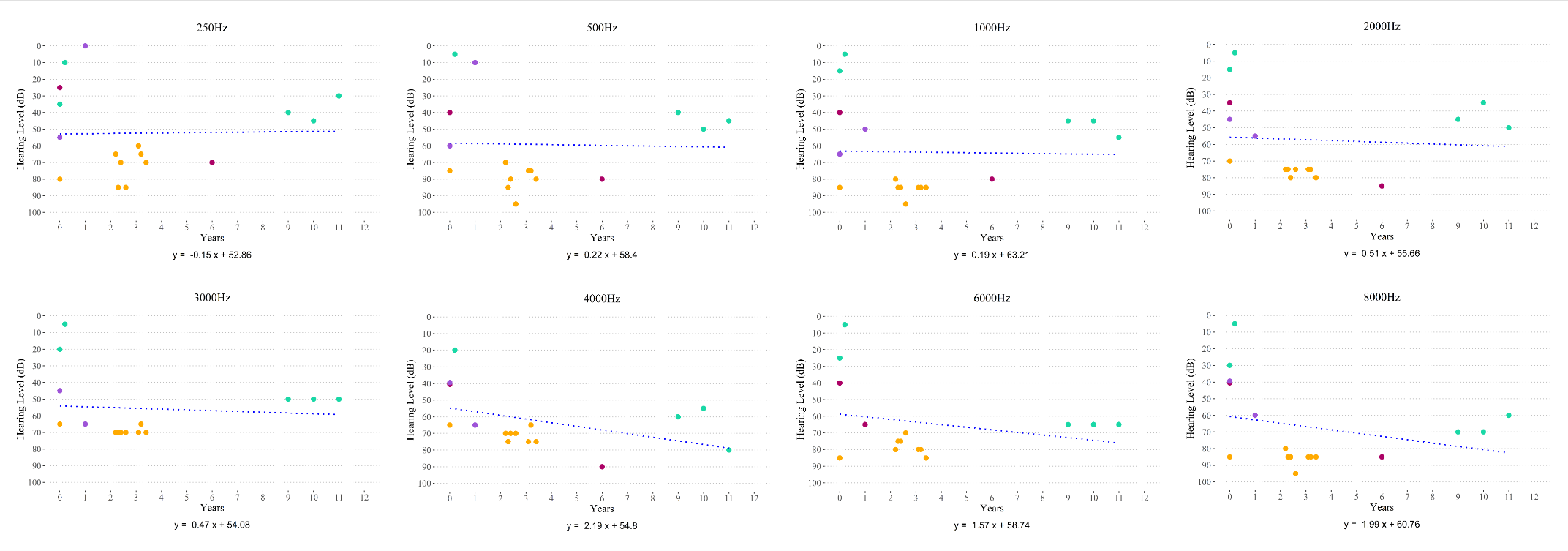

Figure S1. Scattered plot showing hearing thresholds and the duration of the disease for each frequency in the four patients carrying the chr11:17599671C>T variant in *OTOG* gene. Green dots represent the SMD Brazilian proband; Magenta and Violet represent the two SMD Spanish patients; Yellow dots represent the FMD Spanish patient. The worst ear data was used for the plot.
