## Supplementary Figure S2 for "Replication of missense OTOG gene variants in a Brazilian cohort of Menière’s Disease"

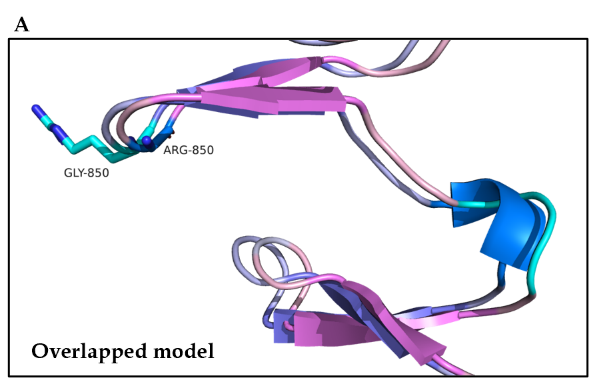

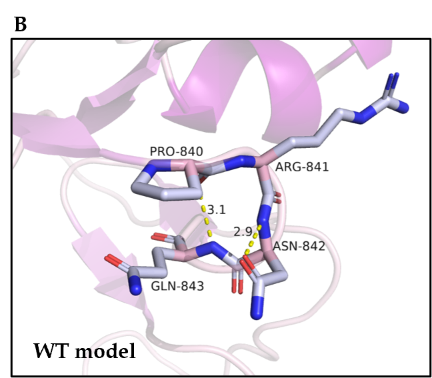

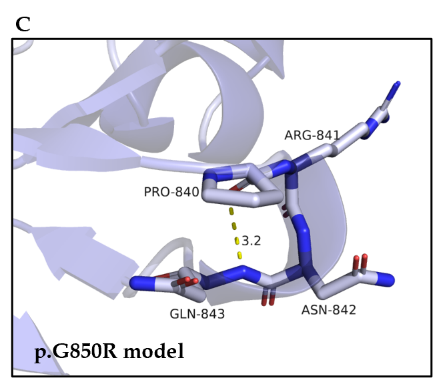

Supplementary Figure S2. Otogelin structure alteration. A. Wild type and mutated Otogelin structures overlapped, showing the creation of a α-helix at p.841-843. B. Wild type Otogelin amino acids arrange and polar contacts at p.840-843. C. Mutated Otogelin amino acids arrange and polar contacts at p.840-843. The polar contact formed within p.Asn842 in the WT structure no longer exists in the mutated one.
