## Supplementary Table S1 for "Replication of missense OTOG gene variants in a Brazilian cohort of Menière’s Disease"

Supplementary Table S1. Coefficient of determination R2 and p value for each scattered plot.

| Frequency (Hz) | p-value | *R^2^* |
| --- | --- | --- |
| 250 | 0.9356 | 0.00045 |
| 500 | 0.9092 | 0.000896 |
| 1000 | 0.9246 | 0.000617 |
| 2000 | 0.7729 | 0.005722 |
| 3000 | 0.7588 | 0.007508 |
| 4000 | 0.0773 | 0.1934 |
| 6000 | 0.9356 | 0.00045 |
| 8000 | 0.2674 | 0.08128 |
