## Supplementary Table S2 for "Replication of missense OTOG gene variants in a Brazilian cohort of Menière’s Disease"

Supplementary Table S2. Human Splicing Finder PRO predictions for chr11:17599671C>T variant in *OTOG* gene.

| Regulatory elements | Name | Position | Sequence | Status |
| --- | --- | --- | --- | --- |
| Exonic Slpicing Enhancer (ESE) | ESE_SRp40 | chr11:17599665 | TTTCAGC | Site Broken |
| Exonic Slpicing Silencer (ESS) | IIE | chr11:17599666 | TTCAGT | Site Created |
| Exonic Slpicing Enhancer (ESE) | EIE | chr11:17599667 | TCAGTG | Site Created |
| Exonic Slpicing Silencer (ESS) | IIE | chr11:17599667 | TCAGTG | Site Created |
| Exonic Slpicing Silencer (ESS) | IIE | chr11:17599668 | CAGTGT | Site Created |
| Exonic Slpicing Enhancer (ESE) | ESE_ASF | chr11:17599668 | CAGCGTA | Site Broken |
| Exonic Slpicing Enhancer (ESE) | ESE_ASFB | chr11:17599668 | CAGCGTA | Site Broken |
| Exonic Slpicing Enhancer (ESE) | ESE_SRp55 | chr11:17599669 | AGCGTA | Site Broken |
| Exonic Slpicing Silencer (ESS) | IIE | chr11:17599669 | AGTGTA | Site Created |
| Exonic Slpicing Silencer (ESS) | Fas ESS | chr11:17599669 | AGTGTA | Site Created |
| Exonic Slpicing Silencer (ESS) | IIE | chr11:17599670 | GTGTAT | Site Created |
| Exonic Slpicing Enhancer (ESE) | RESCUE ESE | chr11:17599671 | CGTATG | Site Broken |
| Exonic Slpicing Enhancer (ESE) | EIE | chr11:17599671 | TGTATG | Site Created |
| Exonic Slpicing Silencer (ESS) | IIE | chr11:17599671 | TGTATG | Site Created |
