## Supplementary Table S3 for "Replication of missense OTOG gene variants in a Brazilian cohort of Menière’s Disease"

Supplementary Table S3. Model quality validation of predicted structural models for selected variants in Otogelin.

| Variant Modelled | Amino acid change | Modelling method | Software used | Evaluation method | | | | |
| --- | --- | --- | --- | --- | --- | --- | --- | --- |
|  |  |  |  | Dope score | molpdf score | GA341 score | ERRAT score | Ramachandran plot |
| chr11:17599671C>T | p.Pro1240Leu | Homology | Modeller v10.5 | -247147.0625 | 18634.0332 | 1.00000 | 75.6295 | 91.9% core |
| chr11:17576581G>T | p.Gly850Arg | Homology | Modeller v10.5 | -247374.6719 | 18558.66992 | 1.00000 | 72.9905 | 91.7% core |
| chr11:17594108 C>T | p.Pro1129Leu | Homology | Modeller v10.5 | -247354.5625 | 18280.60742 | 1.00000 | 73.7158 | 91.7% core |
| chr11:17599671C>T + chr11:17576581G>T + chr11:17594108 C>T | p.Pro1240Leu + p.Gly850Arg + p.Pro1129Leu | Homology | Modeller v10.5 | -246513.3281 | 18042.59961 | 1.00000 | 72.2502 | 91.2% core |
